# The significance of mitochondrial haplogroups in preeclampsia risk

**DOI:** 10.1101/2023.05.22.23290334

**Authors:** Kristina Wendelboe Olsen, Paula L. Hedley, Christian M. Hagen, Line Rode, Sophie Placing, Karen R Wøjdemann, Anne-Cathrine Shalmi, Karin Sundberg, Anne Nørremølle, Ann Tabor, Joanna L. Elson, Michael Christiansen

## Abstract

**Objective:** To determine whether mitochondrial haplogroups function as disease-modifiers or as susceptibility factors in preeclampsia using a traditional haplogroup association model.

**Methods:** This retrospective study haplotyped 235 control and 78 preeclamptic pregnancies from Denmark using either real-time PCR or Sanger sequencing depending on the rarity of the haplogroup.

**Results:** No significant association between haplogroups and the risk of preeclampsia was found, nor was any role for haplogroups in disease severity uncovered.

**Conclusion:** Mitochondrial haplogroups are not associated with preeclampsia or the severity of preeclampsia in the Danish population. However, this study cannot exclude a role for less common mtDNA variation. Models that can examine these should be applied in preeclamptic patients.

**Graphical Abstract:** 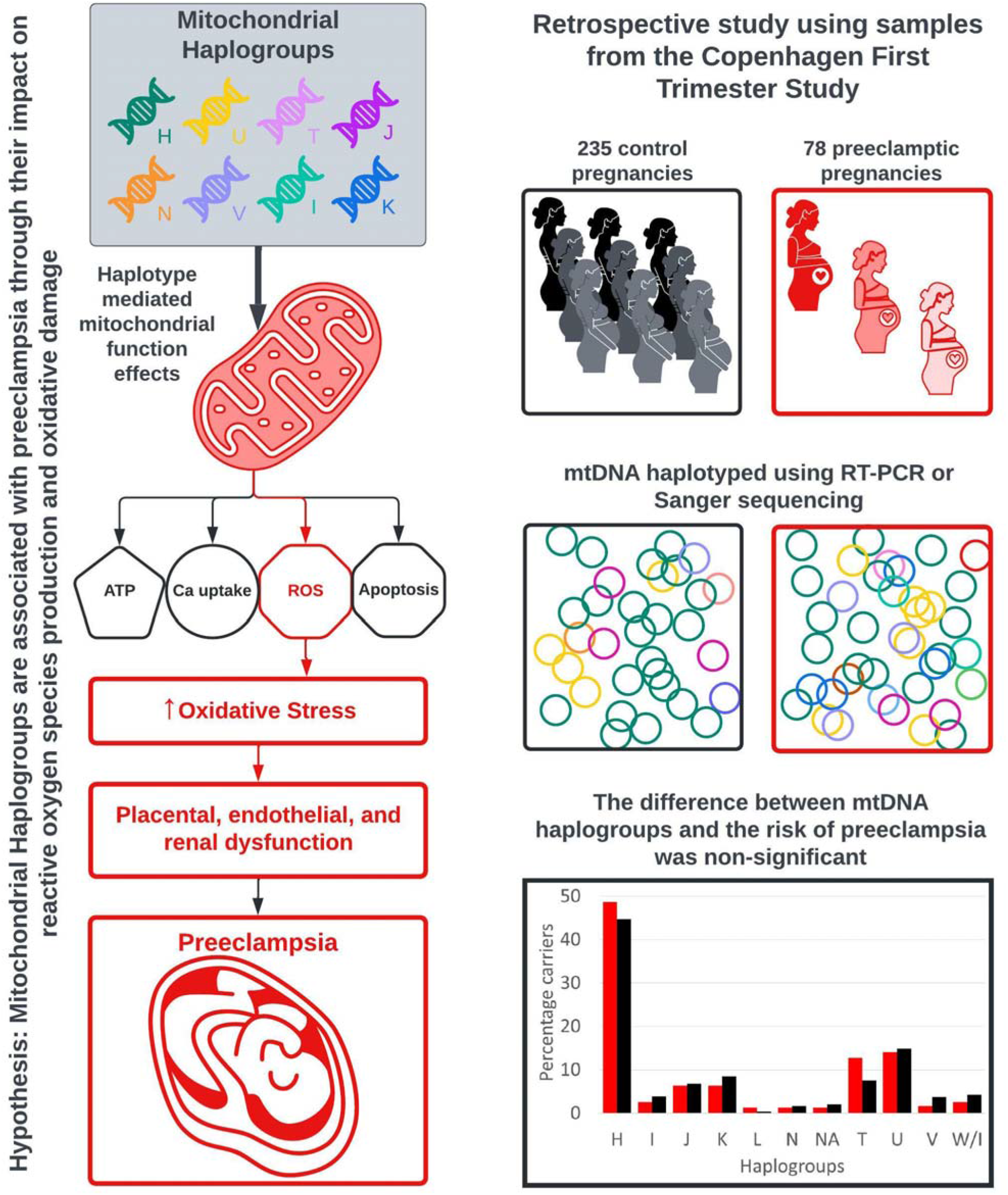

## Introduction

Preeclampsia affects 2-7 % of all pregnancies in the western world, and is the greatest cause of maternal mortality and morbidity worldwide [1-3]. It is a multisystem pregnancy disorder diagnosed by the presence of hypertension and proteinuria with symptoms varying from mild hypertension, proteinuria and edema to multi-organ failure [2, 4, 5]. Preeclampsia has a poorly understood etiology, but it is believed to be influenced by genetic, immunological and environmental factors [6]. Two clinical syndromes results from preeclampsia; one in the mother and one in the fetus [4]. The maternal syndrome is characterized by hypertension, frequently proteinuria, or other systemic abnormalities. In the United Kingdom the main cause of maternal mortality among preeclampsia cases is pulmonary edema [7]. PE is associated with metabolic, cardiac and other disorders later in life for both mother and child [5].

By definition preeclampsia develops after the 20th week and can be classified as early-onset or late-onset; before or after the 34th week of gestation [8]; it can occur with or without intrauterine growth restriction [9] and has been associated with respiratory distress in very low birth weight infants [10]. The risk for adverse neonatal outcome is increased with early-onset preeclampsia [6], which, together with restricted fetal growth is associated with high incidence of stillbirth and perinatal death [11]. Preeclampsia can be divided into two-stage disease: Stage I, the asymptomatic stage with abnormal placentation, and stage II, during which clinical signs and symptoms develop [12, 13]. Preeclampsia is the result of changes initiated in the first trimester of pregnancy [14]. Preventative treatment, with low dose aspirin, started late in the first trimester, has been recommended for high-risk pregnancies [15]. However, the risk of developing preeclampsia is typically – and increasingly - assessed using either demographic variables or clinical history and or biomarkers [16-18].

In healthy pregnancies the placentation begins with the blastocysts adhering to the uterine endometrium (decidua) and forming invasive extravillous cytotrophoblast cells. These cells invade the uterine wall by changing their phenotype from epithelia to endothelia to create vessels for the placental circulation. The predominant theory of the pathogenesis of preeclampsia is that the cytotrophoblasts fail to switch their phenotype, which leads to small, poorly developed spiral arteries resulting in a compromised maternal-fetal blood flow [19]. Persistent defective placental perfusion produces placental hypoxia which will subsequently lead to oxidative stress [20, 21]. Oxidative stress has been shown to be present in placental tissue, moreover there is a higher level of oxidative stress in preeclampsia and other pregnancy-related pathologies [22, 23], indeed, higher levels of oxidative stress have been shown to be present prior to the diagnosis of PE [24]. Furthermore, aberrant placental energy metabolism may explain some of the signs and symptoms of preeclampsia [25], and mitochondrial proteins have also been associated with apoptosis, oxidative stress, reactive oxygen species (ROS) generation and mitochondrial damage [26]. This suggests that mitochondria play a role in the development of preeclampsia.

The mitochondria are responsible for producing the majority of cellular ATP by oxidative phosphorylation which also produces ROS such as superoxide (O2-) and hydrogen peroxide (H2O2). It has been assumed that these ROS are continually generated as a result of mitochondrial respiration and are key to some disease processes as well as ageing [27]. Our understanding of ROS has increased in recent years following the elucidation of their role as signaling molecules [28]. The presence of mitochondrial DNA (mtDNA), the mitochondria’s own genetic material, is unique among cellular organelles. This circular, double stranded molecule encompasses ∼16 kbp of DNA in humans [29]. It codes for 13 polypeptides of the mitochondrial respiratory chain, two ribosomal RNAs and the 22 transfer RNAs necessary for the mitochondrial protein synthesis in the matrix. All other mitochondrial proteins are encoded by nuclear genes. Each mammalian cell contains several hundreds or thousands of mitochondria depending on the cell’s energy demands [30]. MtDNA is maternally inherited which means the evolution of mtDNA is defined by the emergence of distinct lineages called haplogroups [31]. Variation in mtDNA has been linked to a number of complex traits [32, 33] and it has been suggested that this might be linked to differential mitochondrial performance but this is a controversial field [34].

Torbergsen and coworkers were the first to discover a high incidence of preeclampsia in families with mitochondrial dysfunction [35]; an indication that mitochondria are involved in preeclampsia [36, 37] and thus potentially mtDNA variation could affect the function of the placenta, and thereby lead to the development of preeclampsia. The mitochondrial haplogroup H, by far the most frequent in Denmark [38], has been shown by some to have higher VO2max but is also associated with increased mitochondrial oxidative stress compared to some other haplogroups [39, 40]. These studies have been supported by more recent work looking at athletic performance [41], and an association to heart disease [42]. As with many observations in the mitochondrial association field this has been controversial [43]. However, we hypothesized that, if this were true, haplogroup H will be overrepresented in first-trimester pregnancies which would go on to develop preeclampsia compared to non-complicated pregnancies. The idea of comparing haplogroup H to other European groups has been taken due to the high frequency of haplogroup H in the European population [44].

## Materials and Methods

### Study populations

The study cohort comprised of 78 Danish women with preeclampsia and a control group of 235 pregnant Danish women. The study was a case-control sub study, using patients where dried blood spot samples were available, of the Copenhagen First Trimester Screening Study (CFTSS) [45]. Several biomarker studies in PE have been conducted as sub studies of the CFTSS [46-49]. The clinical and demographical data of participants are given in table 1. General population control data were obtained from the Copenhagen City Heart Study (Østerbroundersøgelsen) [50], where the haplotyping is based on haplogroup specific markers in the hypervariable regions (HV1 and HV2) of the mtDNA.

**Table 1.**
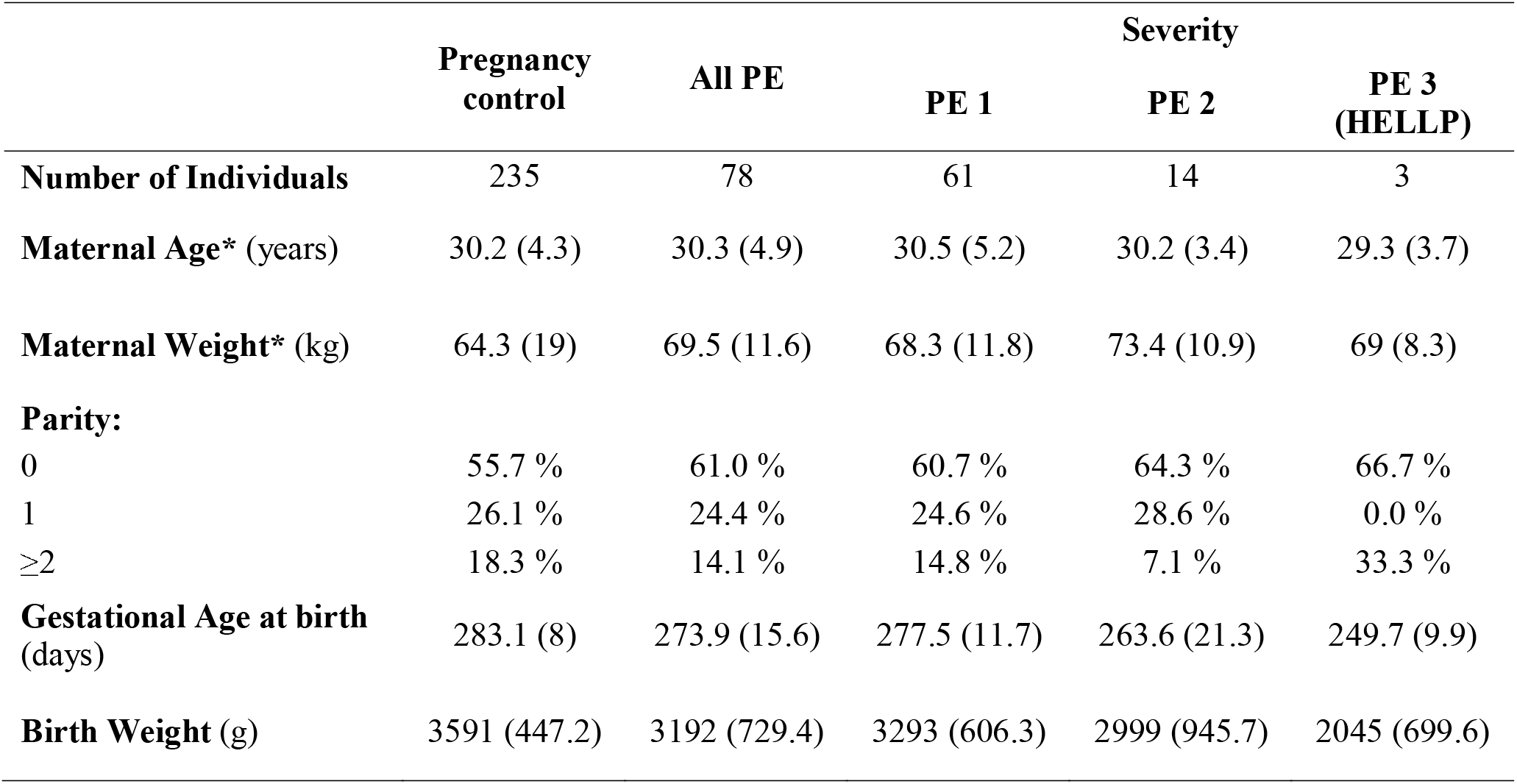
Clinical and demographic data of the preeclampsia (PE) cases and pregnancy controls. ‘Control’ is the group of healthy pregnancies. ‘PE’ is the preeclampsia patients. †Severity refers to the severity of PE (PE 1=uncomplicated PE, PE 2=severe PE, PE 3=HELLP syndrome). Parity refers to the number of times the woman has given birth.

In this study, preeclampsia was defined according to the guidelines of the Danish Society for Obstetrics and Gynecology (www.dsog.dk)[51], except that proteinuria was required. The maternal systolic blood pressure should be more than 140 mmHg and diastolic blood pressure more than 90 mm Hg, along with ≥ 0.3 g protein in a urine specimen after 20 weeks of gestation in women with previously normal blood pressure. Severe PE had BP > 160 mm Hg (sys) and/or 110 mm Hg(dia) and/or severe organ involvement, as defined in the guidelines[51]. HELLP syndrome required laboratory signs of hemolysis, ALAT/ASAT > 70 U/L, and less than 10^9^ platelets per L [51].

### Haplotyping of mtDNA

For the determination of mitochondrial haplogroup all 78 patients and 235 controls were haplotyped. Most of them were haplotyped using TaqMan real time PCR [52], while individuals with an uncommon haplogroup were examined with Sanger Sequencing.

### Statistics

Frequencies of all mitochondrial haplogroups in preeclamptic patients and controls were tested for independence using Fisher’s exact test as appropriate. Similarly, frequencies of mitochondrial haplogroups were compared between controls from a general population study and pregnancy controls. Odds-ratios and 95% confidence intervals were calculated.

### Ethics statement

The Copenhagen First Trimester Screening Study was conducted according to the guidelines of the Declaration of Helsinki and approved by the Scientific ethics committee of the cities of Copenhagen and Frederiksberg (No. (KF) 01-288/97) and the Data protection Agency[45].

## Results

### Haplogroup distribution

The distribution of haplogroups in preeclamptic pregnancies as well as control pregnancies and in a Danish population control group [45, 50] is given in table 2. These results were obtained from the TaqMan real time PCR and the Sanger Sequencing.

**Table 2.** Haplogroup distribution for preeclampsia (PE) pregnancies, control pregnancies and the general population controls from Benn et al. 2008.

| **Haplogroup** | **PE (%)** | **Pregnancy**<br>**Control (%)** | **Population**<br>**Control (%)** |
| --- | --- | --- | --- |
| **A** | 0 | 2 (0.85) | n.d. |
| **B** | 0 | 1 (0.4) | n.d. |
| **C** | 0 | 1 (0.4) | n.d. |
| **H** | 38 (48.7) | 105 (44.7) | 4244 (45.9) |
| **HV** | 0 | 5 (2.1) | 412† (4.5) |
| **I** | 2 (2.6) | 9 (3.9) | 350* (3.8) |
| **J** | 5 (6.4) | 16 (6.8) | 843 (9.1) |
| **K** | 5 (6.4) | 20 (8.5) | 571 (6.2) |
| **L** | 1 (1.3) | 1 (0.4) | n.d. |
| **N** | 1 (1.3) | 4 (1.7) | n.d. |
| **R** | 0 | 1 (0.4) | n.d. |
| **T** | 10 (12.8) | 18 (7.6) | 912 (9.9) |
| **U** | 11 (14.1) | 35 (14.9) | 35 (14.9) |
| **V** | 4 (1.7) | 9 (3.8) | 412† (4.5) |
| **W** | 0 | 1 (0.4) | n.d. |
| **X** | 0 | 1 (0.4) | n.d. |
| **Z** | 0 | 1 (0.4) | 324 (3.5) |
| **W/I** | 2 (2.6) | 10 (4.3) | 350* (3.8) |
| **NA** | 1 (1.3) | 5 (2.1) | n.d. |
| **Total** | 78 (100) | 235 (100) | 9254 (100) |

\* Haplogroup was not distinguished. † haplogroup HV and V were not distinguished. NA not available. n.d. not determined.

### Validation of pregnancy controls

To investigate whether or not the pregnancy controls from CFTSS are representative for the Danish population, Fishers exact test was performed to compare them with the haplogroup distribution in a general Danish population [45], table 3. It shows that there were no significant differences, therefore, it can be concluded that the pregnancy control group is representative for the general Danish population.

**Table 3.** Comparison of the haplogroup distribution in the healthy pregnancy control group and a general Danish population. It shows that there were no significant differences (p-value > 0.05), it can be concluded that the healthy pregnancy control group is representative for the general Danish population.

| Test setup | Pregnancy control/<br>population control,<br>percentage | Fisher's exact<br>test, p-value | Odds-ratio | 95% CI |
| --- | --- | --- | --- | --- |
| **H vs. non-H** | 44.5/46.6 | 0.55 | 1.08 | 0.82-1.43 |
| **U vs. non-U** | 14.9/15.9 | 0.65 | 1.09 | 0.76-1.6 |
| **K vs. non-K** | 8.5/6.2 | 0.21 | 0.72 | 0.42-1.23 |
| **J vs. non-J** | 6.8/9.1 | 0.13 | 1.54 | 0.9-2.82 |
| **T vs. non-T** | 7.6/9.9 | 0.49 | 1.23 | 0.75-2.14 |
| **V vs. non-V** | 3.8/4.5 | 0.79 | 0.63 | 0.63-2.79 |

### Haplogroup distribution and preeclampsia and severity of preeclampsia

The data did not support a link between European haplogroups and PE, table 4. None of the uncorrected p-values indicate significance when comparing the control pregnancies with the PE pregnancies, with the value for the most common of the European haplogroups H being p=0.55. Note that there was no difference when the PE pregnancies were compared to the general Danish population.

**Table 4.** Comparison of preeclamptic pregnancies and healthy control pregnancies for each haplogroup.

| Test setup | PE/<br>pregnancy control,<br>percentage | Fisher's exact<br>test, p-value | Odds-ratio | 95% CI |
| --- | --- | --- | --- | --- |
| **H vs. non-H** | 49/44 | 0.6 | 1.19 | 0.68-2 |
| **U vs. non-U** | 14/15 | 1 | 0.9 | 0.4-2 |
| **K vs. non-K** | 6/8 | 0.64 | 0.73 | 0.2-2.1 |
| **J vs. non-J** | 6/7 | 1 | 0.93 | 0.26-2.8 |
| **T vs. non-T** | 13/8 | 0.17 | 1.78 | 0.7-4.29 |
| **I vs. non-I** | 3/4 | 1 | 0.7 | 0.06-3.3 |
| **V vs. non-V** | 5/4 | 0.74 | 1.4 | 0.3-5 |
| **L vs. non-L** | 1/0.5 | 0.44 | 3 | 0.38-240 |
| **N vs. non-N** | 1/2 | 1 | 0.75 | 0.15-7.76 |

CI: confidence interval

**Table 5.** Comparison of preeclamptic pregnancies and a general population control from Benn et al 2008 for each haplogroup.

| Test setup | PE/<br>Population control,<br>percentage | Fisher's exact<br>test, p-value | Odds-ratio | 95% CI |
| --- | --- | --- | --- | --- |
| **H vs. non-H** | 49/46,6 | 0.73 | 1.09 | 0.67-1.74 |
| **U vs. non-U** | 14/15.9 | 0.64 | 0.83 | 0.4-1.6 |
| **K vs. non-K** | 6/6.2 | 1 | 1.01 | 0.3-2.5 |
| **J vs. non-J** | 6/9.1 | 0.44 | 1.44 | 0.66-2.85 |
| **T vs. non-T** | 13/9.9 | 0.32 | 1.78 | 0.7-4.29 |
| **V vs. non-V** | 5/4.5 | 0.79 | 1.09 | 0.3-3 |

CI: confidence interval

The PE cases were divided into mild (PE 1), severe (PE 2), and HELLP cases (PE 3), Table 1. The distribution of clinical severity as a function of mtDNA haplogroups is depicted in Figure 1. No difference was seen; however, the statistical power is very limited.

**Figure 1.**
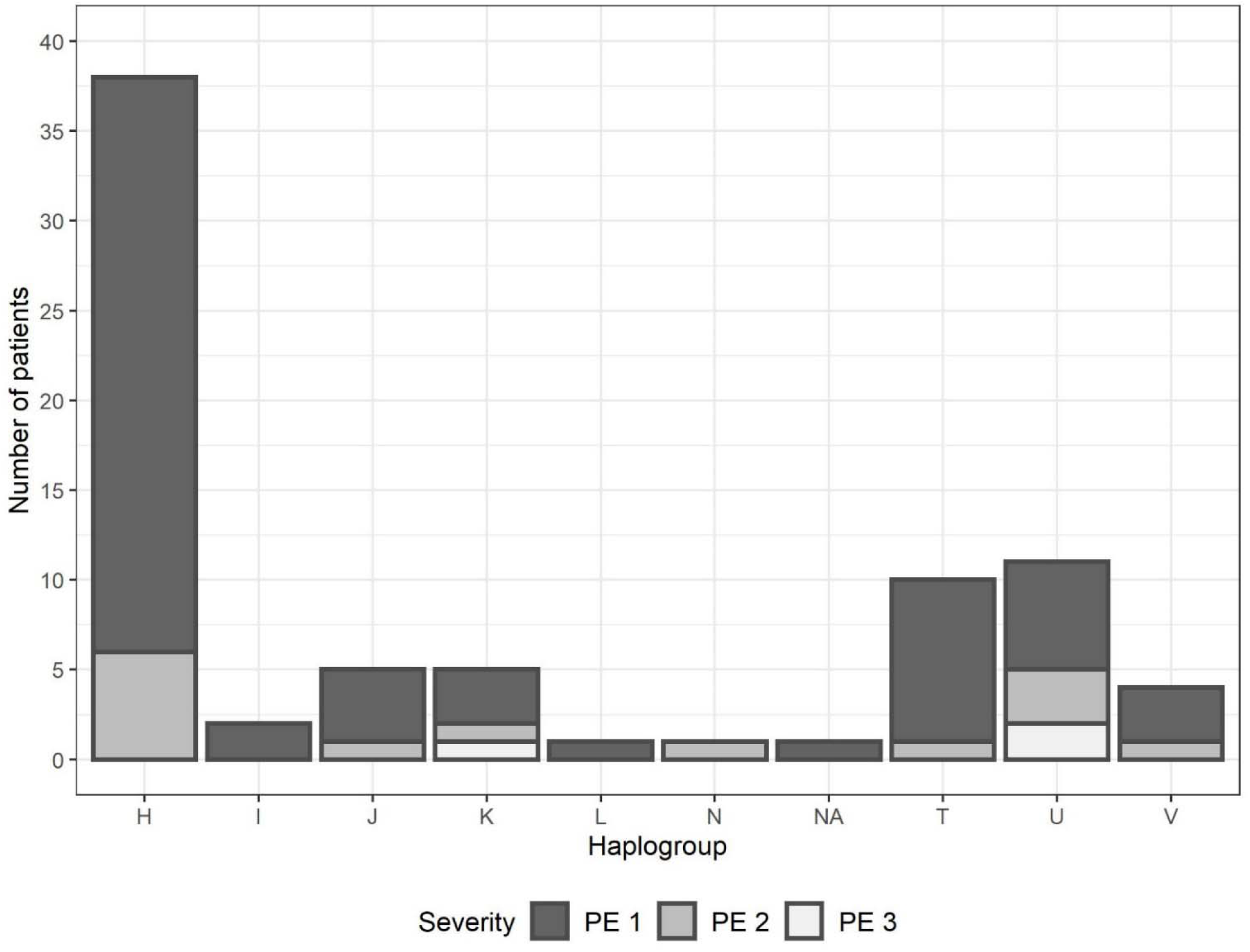
Haplogroup distribution and severity of PE. The x-axis shows the haplogroups, while the y-axis is the number of PE patients. PE 1 is the mildest, uncomplicated form of preeclampsia (PE 1 n=61). PE 2 is severe preeclampsia (PE 2 n=14). PE 3 is preeclampsia complicated with HELLP syndrome (PE 3 n=3). NA is the individuals with an unknown haplogroup.

## Discussion

Several health and disease states have been found to be associated with the different mitochondrial haplogroups [33, 42]. In this study the association between the mitochondrial haplogroups and the development of preeclampsia was studied in an attempt to determine if mitochondrial haplogroups function as disease-modifiers or as susceptibility factors in preeclampsia using a traditional haplogroup association model.

The placenta plays an important role in development of preeclampsia [19, 36]. As mitochondria are maternally inherited [53], the fetus, the placenta and the mother have the same mitochondrial haplogroup; consequently, the placental mtDNA haplogroup can be determined from a blood sample from either the mother or – in retrospective studies – from the child.

Blood and clinical data from 78 patients with preeclampsia and 235 healthy pregnancy controls were employed to investigate the possibility that preeclampsia is associated with haplogroup H as such an association has been demonstrated in Danish hypertrophic cardiomyopathy patients [42]. Since haplogroup H exhibits increased ROS production, oxidative damage and higher VO2max [39, 40] the preeclamptic women are hypothesized to be more likely to have haplogroup H compared to the controls due to the association between preeclampsia and oxidative stress [21, 24].

The CFTSS pregnancy controls were validated by comparing them with the haplogroup distribution from a Danish general population study. This comparison showed no significant differences (p-value between 0.13-0.79 in a Fisher’s exact test), thus the controls fit the distribution seen in the general population, table 2. The haplogroup distribution of preeclamptic patients and healthy controls was compared with Fisher’s exact test. No significant difference in the haplogroup distribution between the different groups (P = 0.98) was found, table 3. When haplogroup H was compared with non-H haplogroups there was likewise no significant difference (P = 0.60). The same is true for the other haplogroups with p-values > 0.05, table 3.

It has also been suggested that mtDNA haplogroups might affect the course of disease [54]. The severity of preeclampsia might be associated with different haplogroups. With more cases of severe preeclampsia, it is possible that mitochondrial haplogroups could be proven to predict the severity/phenotype of the disease. The same might be true for early-onset versus late-onset preeclampsia, since many researchers suggest that they follow two separate pathogenic pathways [55, 56], they might also be associated with different mitochondrial haplogroups. However, our data on severity does not support that this should be the case. And the use of a small number of patients makes the time of birth largely determined by clinical assessment and not biology. It can be difficult to collect data from severe preeclamptic patients, since almost all women in Denmark make use of the offer of free medical care and therefore are often treated before the disease develops to more severe stages. Thus, there might be an undetected association.

Haplogroup H was not found to be associated with preeclampsia, but that does not exclude an association with some of the other haplogroups instead. Because haplogroup J, U, T and K function as disease-modifiers in other diseases they could be the new association candidates. However, to get enough statistical power a larger cohort will be needed or another population with a different haplogroup distribution; a population with high proportion of J, U, T or K. In larger association studies involving rarer mtDNA variants or rare haplogroups it will, however, be very important to account for nuclear genomic differences [57].

The placental mtDNA content (mtDNA copy number) has shown to be a molecular marker of mitochondrial damage and mitochondrial inflammation [58]. Exposure to particulate matter in third trimester has been shown to be associated with mitochondrial damage exemplified by a lower mtDNA content [58]. A significant decrease in mtDNA content is also observed in placenta from smokers compared with non-smokers [59]. Since environmental factors show this impact on mitochondria, it might be that the same can be seen in preeclampsia, only with lower mtDNA content already in the first trimester where the placenta is poorly implanted. The dysfunctional placental mitochondria in preeclampsia might be due to a small copy number of mtDNA in the first trimester of pregnancy [60, 61].

## Conclusion

A significant association between mtDNA haplogroups and the risk of preeclampsia was not demonstrated. However, the diversity of the haplogroups found makes it possible that haplogroups with low prevalence may actually play a role for occurrence or clinical presentation of PE. There is currently no basis for considering mtDNA haplogroups potential first trimester risk markers of PE.

## Data Availability

All data produced in the present work are contained in the manuscript

## Acknowledgements

We would like to acknowledge the technical assistance of Dennis Jelsbak Schmidt. This research has been conducted using the Danish National Biobank resource, supported by the Novo Nordisk Foundation.

